# Systematic review and meta-analysis of predictive symptoms and comorbidities for severe COVID-19 infection

**DOI:** 10.1101/2020.03.15.20035360

**Authors:** V Jain, J-M Yuan

**Affiliations:** Institute for Global Health, University College London (UCL), UK; Public Health, London Borough of Hackney, London, UK; Public Health, London Boroughs of Camden & Islington, London, UK

## Abstract

**Background/introduction:** COVID-19, a novel coronavirus outbreak starting in China, is now a rapidly developing public health emergency of international concern. The clinical spectrum of COVID-19 disease is varied, and identifying factors associated with severe disease has been described as an urgent research priority. It has been noted that elderly patients with pre-existing comorbidities are more vulnerable to more severe disease. However, the specific symptoms and comorbidities that most strongly predict disease severity are unclear. We performed a systematic review and meta-analysis to identify the symptoms and comorbidities predictive of COVID-19 severity.

**Method:** This study was prospectively registered on PROSPERO. A literature search was performed in three databases (MEDLINE, EMBASE and Global Health) for studies indexed up to 5^th^March 2020. Two reviewers independently screened the literature and both also completed data extraction. Quality appraisal of studies was performed using the STROBE checklist. Random effects meta-analysis was performed for selected symptoms and comorbidities to identify those most associated with severe COVID-19 infection or ICU admission.

**Results:** Of the 2259 studies identified, 42 were selected after title and abstract analysis, and 7 studies (including 1813 COVID-19 patients) were chosen for inclusion. The ICU group were older (62.4 years) compared to the non-ICU group (46 years), with a significantly higher proportion of males (67.2% vs. 57.1%, p=0.04). Dyspnoea was the only significant symptom predictive for both severe disease (pOR 3.70, 95% CI 1.83 – 7.46) and ICU admission (pOR 6.55, 95% CI 4.28– 10.0). Notwithstanding the low prevalence of COPD in severe disease and ICU-admitted groups (4.5% and 9.7%, respectively), COPD was the most strongly predictive comorbidity for both severe disease (pOR 6.42, 95% CI 2.44 – 16.9) and ICU admission (pOR 17.8, 95% CI 6.56 – 48.2). Cardiovascular disease and hypertension were also strongly predictive for both severe disease and ICU admission. Those with CVD and hypertension were 4.4 (95% CI 2.64 – 7.47) and 3.7 (95% CI 2.22 – 5.99) times more likely to have an ICU admission respectively, compared to patients without the comorbidity.

**Conclusions:** Dyspnoea was the only symptom strongly predictive for both severe disease and ICU admission, and could be useful in guiding clinical management decisions early in the course of illness. When looking at ICU-admitted patients, who represent the more severe end of the spectrum of clinical severity, COPD patients are particularly vulnerable, and those with cardiovascular disease and hypertension are also at a high-risk of severe illness. To aid clinical assessment, risk stratification, efficient resource allocation, and targeted public health interventions, future research must aim to further define those at high-risk of severe illness with COVID-19.

## Introduction

The ongoing novel coronavirus (COVID-19) outbreak is a public health emergency of international concern (PHEIC), involving a novel type of coronavirus originally identified in Wuhan, China. At the time of writing, there have been 105,586 confirmed cases around the world with 3,584 deaths (1). Defining the spectrum of clinical manifestations and the risk factors for severe COVID-19 infections has been identified as an urgent research priority (2,3).

As the virus spreads globally it is likely that government strategies will shift from containment and delay towards mitigation (4). This will involve rapidly scaling up healthcare resources including staff, equipment, facilities, and training, to effectively identify and treat patients. To maximise the use of these limited resources it will be imperative that clinicians are able to triage COVID-19 patients likely to recover after a mild illness from those who are not. In order to do this, a better understanding of the symptoms and comorbidities (which are the first and most routinely collected components of patient data) related to COVID-19 severity is required. This can improve patient outcomes through three chief mechanisms: early clinical intervention in high-risk patients, designing appropriate clinical pathways and risk prediction tools, and the efficient allocation of scarce resources and expensive treatments. Further still, the early identification of individuals more likely to deteriorate can help direct appropriate public health actions to protect the vulnerable and prevent further spread of infection.

A recent meta-analysis of symptoms in 50,466 COVID-19 patients from 10 studies found that fever and cough were the most common symptoms, with 89.1% and 72.2% experiencing these, respectively (5). It also found that the case fatality rate (CFR) was 4.3%, but the association between individual patient factors and severe infection was not investigated. Most reported cases have occurred in adults (median age 59 years) (6). According to most recent US Centers for Disease Control and Prevention (CDC) guidance, risk factors for severe illness are not yet clear, although older patients and those with chronic medical conditions may be at higher risk. (7) The primary aim of this study is therefore to conduct a systematic review and meta-analysis, aggregating all currently available data from published studies, of symptoms and comorbidities predictive for severe illness with COVID-19.

## Methodology

### Retrieval of studies

This study was prospectively registered on PROSPERO. Identification of relevant existing literature was performed by an online search in three databases: MEDLINE, EMBASE and Global Health, for studies published from 1^st^January 2019 to 5^th^March 2020. The MESH headings (keywords) searched were ‘nCoV*’ or ‘coronavirus’ or ‘SARS-2-CoV’ or ‘COVID*’ and ‘symptom*’ or ‘clinical’ or ‘predict*’ or ‘characteristic*’ or ‘co-morbidit*’ or ‘comorbidit*’ or ‘condition*’. Two reviewers (VG, JMY) independently screened the list of titles and abstracts, and the full text of chosen manuscripts. Disagreements on which manuscripts to include during both title and abstract screen, and the subsequent full-text analysis, were discussed until a conclusion was reached. In addition to the MEDLINE/EMBASE/Global Health search, citation tracking was used to identify any remaining relevant published studies, though none were identified. Unpublished studies were not retrieved due to uncertain data quality.

### Inclusion and exclusion criteria

All studies evaluating individual symptoms and comorbidities in predicting severe infection (as measured by disease severity criteria, or ICU admission) were included. All studies of any design, from any time since the outbreak started (in December 2019) were eligible, except case reports of individual patients or literature reviews. To avoid selection bias, no subjective quality criteria were applied to the studies for inclusion. Exclusion criteria included: [1] studies of exclusively paediatric or pregnant patients, due to the varying presentation of COVID-19 in these groups, [2] insufficient data on symptoms/comorbidities on admission in either severe or non-severe disease groups (or ICU and non-ICU groups), [3] coronavirus strains other than COVID-19 and [4] studies not written in English, because of practical limitations with translation.

### Data Extraction

Two reviewers independently extracted data from the included studies for both narrative synthesis and statistical analysis. From each study, various details including the study population, investigated predictive symptoms or comorbidities, and the definitions used to measure outcomes, were extracted into Microsoft excel. These details are presented by study in Table 1. The number of patients in each study, both with and without each symptom or comorbidity, was extracted for statistical analysis (described below).

**Table 1.** Studies investigating the predictive value of symptoms for disease severity or ICU admission

| Study | Year and location | Design | Population (n) | Median age (IQR) | Severe COVID-19 cases (N) | Number of symptoms (comorbidities) investigated | Duration of symptoms before admission (median number of days, IQR) | Definition of outcome measure | Key reported findings relating to COVID-19 severity/ICU admission | Quality of Study* |
| --- | --- | --- | --- | --- | --- | --- | --- | --- | --- | --- |
| Guan et al(10) | December 11, 2019 – January 29, 2020, 522 hospitals from 30 provinces in China | Retrospective Multi-Centre Cohort | n=1099, 58.1% male | 47 (35-58) | N = 67 primary end-point<br>N = 173 severe | 14 (9) | N/A | Composite endpoint: ICU admission, use of mechanical ventilation or death<br>Severity defined as per American Thoracic Society guidelines for community-acquired pneumonia | Patients with severe disease were older than those with non-severe disease by a median of 7 years. Comorbidities were also more common among patients with severe disease (38.7% vs. 21.0%) including COPD (3.5% vs. 0.6%), Diabetes (16.2% vs. 5.7%), hypertension (23.7% vs. 13.4%), and CHD (5.8% vs. 1.8%). | + |
| Huang et al(9) | December 16, 2019 – January 2, 2020, Wuhan, China | Retrospective Multi-Centre Cohort | n=41, 73% male | 49 (41-58) | N = 13 | 8 (6) | ICU care = 7 (4 – 8)<br>No ICU care = 7 (4 – 8.5) | ICU admission | Dyspnoea was significantly more common in the ICU care group (92% vs. 37%, p = 0.001). The presence of various comorbidities was similar across both groups. | ++ |
| Li et al(8) | January – February 2020, 3 hospitals, China | Retrospective Multi-Centre Cohort | N=83, 53% male | Mean age = 45.5 (SD 12.3) | N = 25 | 8 (4) | Severe/critical = 8 (6 – 12)<br>Non-severe/critical = 6 (3 – 8.5) | Severe/critical patients = any of respiratory distress with RR ≥30 breaths per minute, oxygen saturation ≤93%, PaO<sub>2</sub>/FiO<sub>2</sub> ≤300mmHg, required mechanical ventilation, shock occurred, or other organ failure needing ICU care | On univariable logistic regression, factors significantly more predictive for severe/critical cases were age >50 (OR 7.60, 95% CI 2.6 – 21.7), comorbidities (OR 10.6, 95% CI 2.93 – 38.4), dyspnoea (OR 10.9, 95% CI 2.07 – 57.2), chest pain (OR 10.9, 95% CI 1.15 – 102.8), cough (OR 9.95, 95% CI 1.25 – 79.6), and expectoration (OR 4.88, 95% CI 1.51 – 15.8). | + |
| Tian et al(13) | Jan 20 – Feb 10 2020, Beijing, China | Retrospective Multi-Centre Cohort | n=262, 48.5% male | 47.5 (range = 1 – 94) | N = 46 | 5 (0) | Severe = 5.2 (range 0.6 – 9.8)<br>Non-severe = 4.4 (range 0.9 – 7.9) | Mild case = confirmed case with fever, respiratory symptoms and radiographic evidence of pneumonia<br>Severe case = mild case with dyspnoea or respiratory failure | Dyspnoea was the only symptom found to be significantly more common in severe cases compared to non-severe cases (32.6% vs. 1.4%, p<0.001). | + |
| Wang et al(11) | January 1 - 28 2020, Zhongnan Hospital, Wuhan, China | Retrospective Single-Centre Cohort | n=138, 54.3% male | 65 (IQR 42 – 68) | N = 36 | 14 (9) | ICU = 8 (4.5-10)<br>Non-ICU = 6 (3-7) | ICU admission | ICU patients (n = 36), compared with patients not treated in the ICU (n = 102), were older (median age, 66 years vs. 51 years), were more likely to have underlying comorbidities (26 [72.2%] vs. 38 [37.3%]), and were more likely to have dyspnoea (23 [63.9%] vs. 20 [19.6%]), and anorexia (24 [66.7%] vs. 31 [30.4%]). | ++ |
| Xu et al(14) | January – February 2020, China | Retrospective Single-Centre Cohort | n=50, 58% male | 10 % <18 years, 60% 18-50 years, 30% >50 years | N = 13 | 8 (0) | N/A | Severe case = respiratory distress with RR>30, SpO2<93% or PaO2/FiO2<300mmHg<br>Critical case = respiratory failure needing mechanical ventilation, shock, or combination with other organ failure needing ICU care | The most common symptoms were mild fever (37.3C – 38C) in 44% of all cases (51% vs. 23% in severe/critical vs. mild/moderate groups), and cough in 40% of all cases (46% vs. 38% for severe/critical vs. mild/moderate). | - |
| Zhang et al(12) | January 16 – February 3 2020, No. 7 Hospital of Wuhan, China | Retrospective Single-Centre Cohort | n=140, 50.7% male | 57 (range 25-87) | N = 58 | 10 (22) | Severe = 7 (6-12)<br>Non-severe = 8 (5 – 11) | Severe = respiratory distress with RR≥30, SpO2≤93% or PaO2/FiO2≤300 | Having any comorbidity was more common in severe disease patients compared to non-severe (79.3% vs. 53.7%, p = 0.002). Cough (84.9% vs. 67.2%, p=0.02) was significantly more common in the severe group, and nausea significantly less common, compared with the non-severe group ((8.8% vs. 23.2, p=0.02). | + |

\*Quality of Included Studies (% of STROBE checklist criteria met, <55% = -, 55-65% = +, >65% = ++)

### Predictors and Outcomes

The symptoms or comorbidities presented were investigated in at least three included studies. Where studies measured symptoms ambiguously (including abdominal pain/diarrhea(8), myalgia/fatigue (9), and nausea/vomiting (10)), this data was excluded. Some studies reported heart disease and stroke separately (10-12). To allow comparability between studies for meta-analysis, these were grouped into a single predictor (cardiovascular disease). One study was excluded from the analysis of dyspnoea as a predictor of severity, as dyspnoea was part of the definition for severity used by the authors (13).

For disease severity, the included studies varied in their differentiation of patients’ disease status, with classifications of ‘mild, moderate, severe and critical’ (14), ‘ordinary and severe/critical’ (8), ‘common and severe’ (13), and ‘non-severe and severe’, disease (10,12). The first outcome measure used was severe (including both severe and critical cases) vs. non-severe disease. For ICU admission, the included studies varied in their definition of ICU admission, with classifications of ‘ICU, mechanical ventilation or death and non-ICU’ (10), and ‘ICU and non-ICU’ (9,11). The second outcome measure used was ICU admission (including ICU, mechanical ventilation or death, where data was grouped together) vs. non-ICU admission.

### Statistical Analysis

Patient numbers were aggregated across all included studies for each group included in the meta-analysis. Gender was compared between groups using the Chi2 test in STATA (15). This was not possible for age due to a lack of individual-level data. The predictive value of symptoms and comorbidities for each of severe disease and ICU admission was estimated with random effects meta-analysis in STATA. Random effects models were used to account for between study heterogeneity (16), which was estimated with Tau-squared. This provided a pooled odds ratio (pOR), 95% confidence intervals, and a p-value, for each symptom or comorbidity. A p-value of <0.05 was used as a marker for evidence of significant association. Detailed forest plots of the predictive symptoms and comorbidities common to both disease severity and ICU admission are illustrated in the supplementary information file

## Results

The PRISMA flow diagram (Figure 1) illustrates the process for selection of papers in this study.

**Figure 1.**
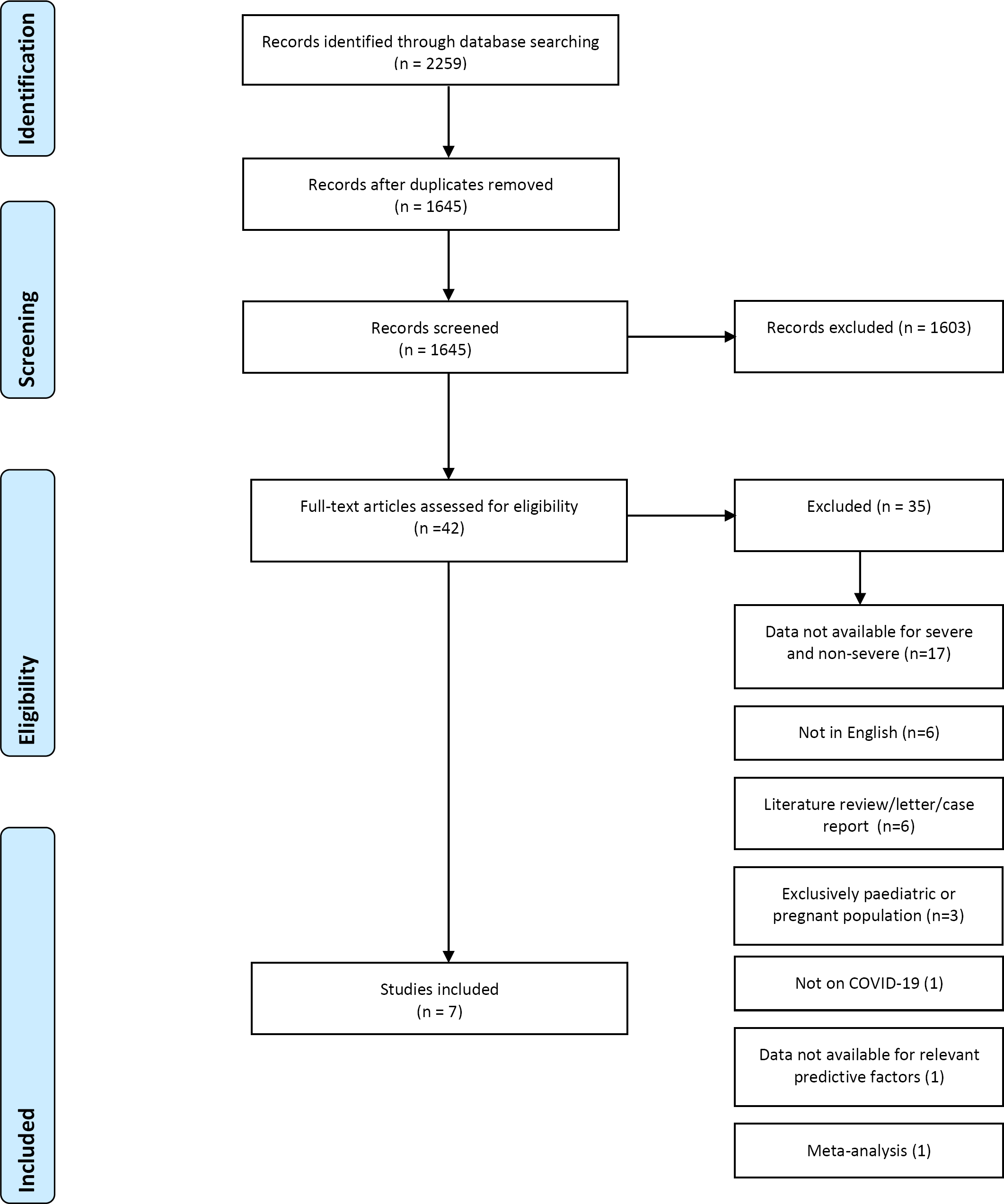
PRISMA Flow Diagram – Included Studies.

### Literature search

The initial search on MEDLINE, EMBASE and Global Health produced 2259 results. After removing duplicates and applying exclusion criteria, there were 42 papers meeting our criteria from title and abstract analysis. On further review, the majority of these studies did not compare proportions of patients with symptoms or comorbidities between severe (or ICU admitted) and non-severe disease (or non-ICU admitted) groups. The reasons for all study exclusions are outlined in Figure 1. A total of seven studies were selected for inclusion.

### Description of Included Studies

Table 1 shows details of all included studies including reported findings pertaining to symptoms and comorbidities related to disease severity or ICU admission. These 7 studies reported on a total of 1813 patients. All included studies were retrospective cohort studies in design, conducted between December 2019 and February 2020 in China, during the novel coronavirus (SARS-2-CoV) outbreak. Guan et al (10) conducted the largest study with 1099 COVID-19 patients, whilst Huang et al (9) included 40 patients in their study. The number of symptoms investigated varied from 5 in one study (13) to 14 in others (10,11). The range of comorbidities investigated varied greatly with two studies not including any (13,14) and one including 22 comorbidities (12).

Table 2 displays the median age and gender of severe and non-severe disease, and ICU and non-ICU admitted, patients, after aggregating all studies. The median age was 62.4 years for ICU admitted patients compared to 46 years for non-ICU patients, and 49.4 years for severe compared to 41.7 years for non-severe disease patients.

**Table 2.**
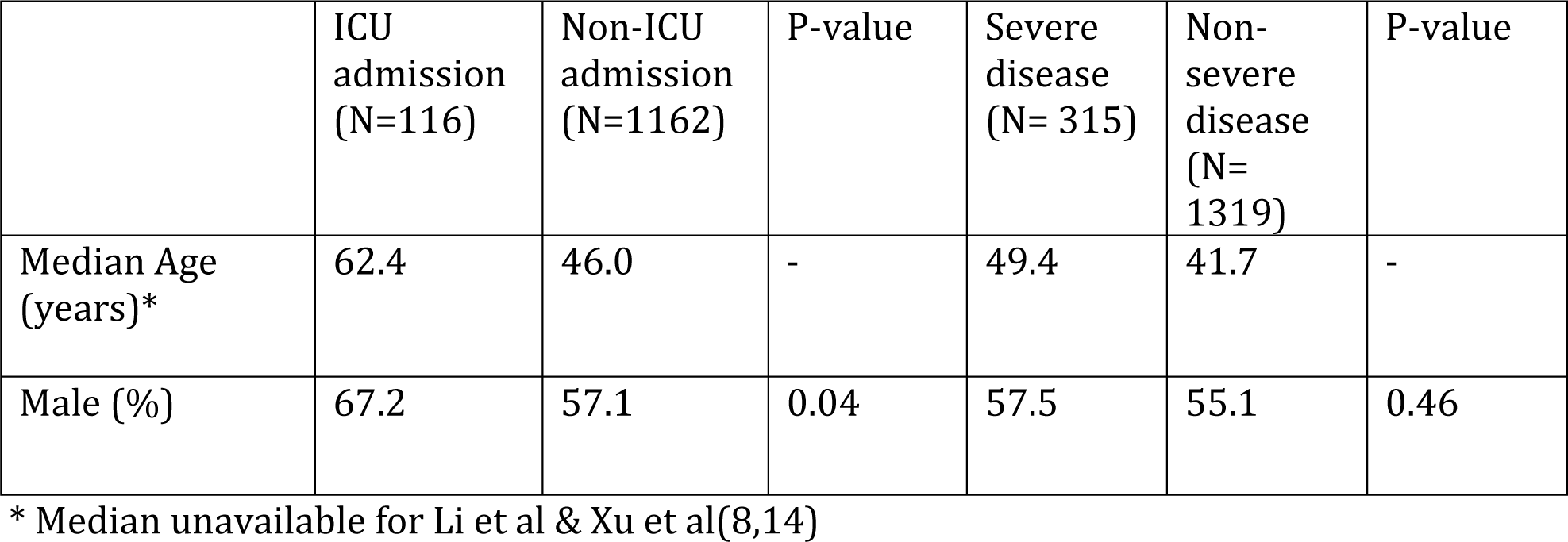
Total Population from Included Studies

### Quality of Included Studies

All 7 included studies were retrospective cohort studies, and were critically appraised using the STROBE checklist (17). The 22 items on the STROBE checklist were formulated into 47 individual indicators, against which each study was marked. Figure 2 illustrates the proportion of included studies which met each individual appraisal indicator. Each paper was assigned an overall quality score based on the percentage of STROBE checklist criteria met (<55% = -, 55-65% = +, >65% = ++), as outlined in Table 1.

**Figure 2.**
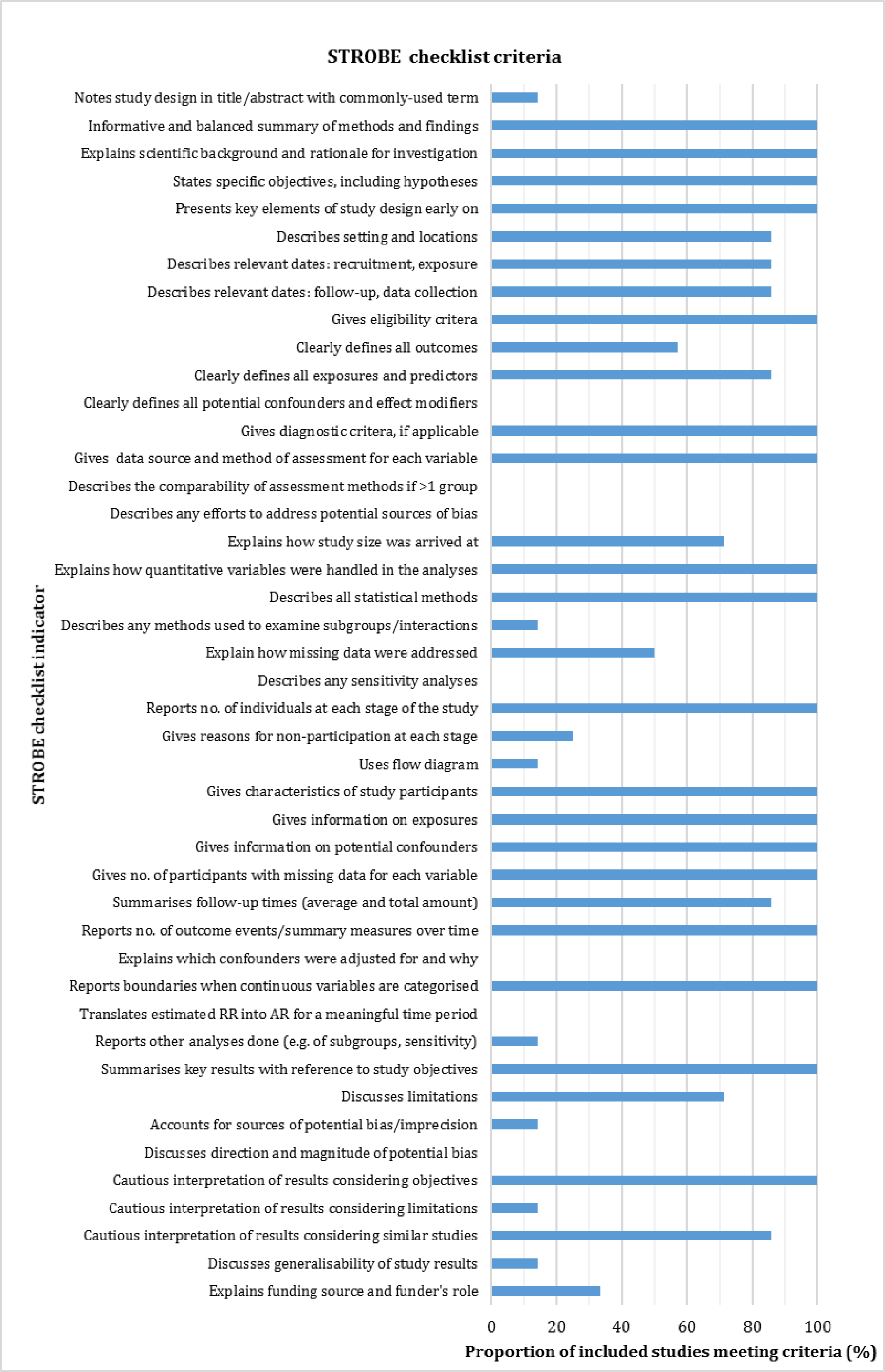
Critical Appraisal of Included Studies.

Appraising with the STROBE checklist highlighted several major weaknesses in the included studies. Firstly, there was no consistent definition on what constituted severe disease. The WHO-China Joint Mission on COVID-19 (18) defined a severe case as tachypnoea (≥30 breaths/min) or oxygen saturation ≤93% at rest, or PaO2/FiO2<300mmHg. Critical cases were defined as respiratory failure requiring mechanical ventilation, shock, or other organ failure that requires intensive care. Although the above criteria were used in some included studies, many defined only one out of severe and critical, with one (8) using a definition for a single severe/critical cohort. One study reported both severity and critical cases (10), using criteria set by the American Thoracic Society to judge severity. Secondly, the time at which severity of disease was determined was not always clear. Severity was assessed on admission in two studies (10,12), whilst three studies did not specify when severity was assessed (8,13,14). It is possible, therefore, that the non-severe group included patients who went on to later develop severe disease. Thirdly, the time point at which symptoms were measured varied from illness onset (via recall) (9,11-13) to clinical presentation (10,14). In the study by Li et al, it was not clear when symptoms were measured (8). Finally, no study specified how each individual symptom or comorbidity was measured. For instance, it was unclear whether fever was objectively measured, and if so, how or by whom. Studies may therefore have been susceptible to measurement and reporting bias.

### Meta-analysis

Tables 3 and 4 show the odds ratios, 95% confidence intervals and p-values for the individual symptoms and comorbidities that were investigated in at least three of the included studies, for both severe disease and ICU admission, respectively. A total of seven symptoms were included in the model for severe disease and six for ICU admission, as well as four comorbidities in both. The most prevalent symptoms in the severe group were cough (70.5%), fever (64.1%) and fatigue (44.5%); in the ICU group these were cough (67.2%), fever (62.9%) and dyspnoea (61.2%). The most prevalent comorbidities in the severe group were hypertension (25.4%) and diabetes (16.8%) and in the ICU group were hypertension (40.5%) and CVD (24.1%).

**Table 3.**
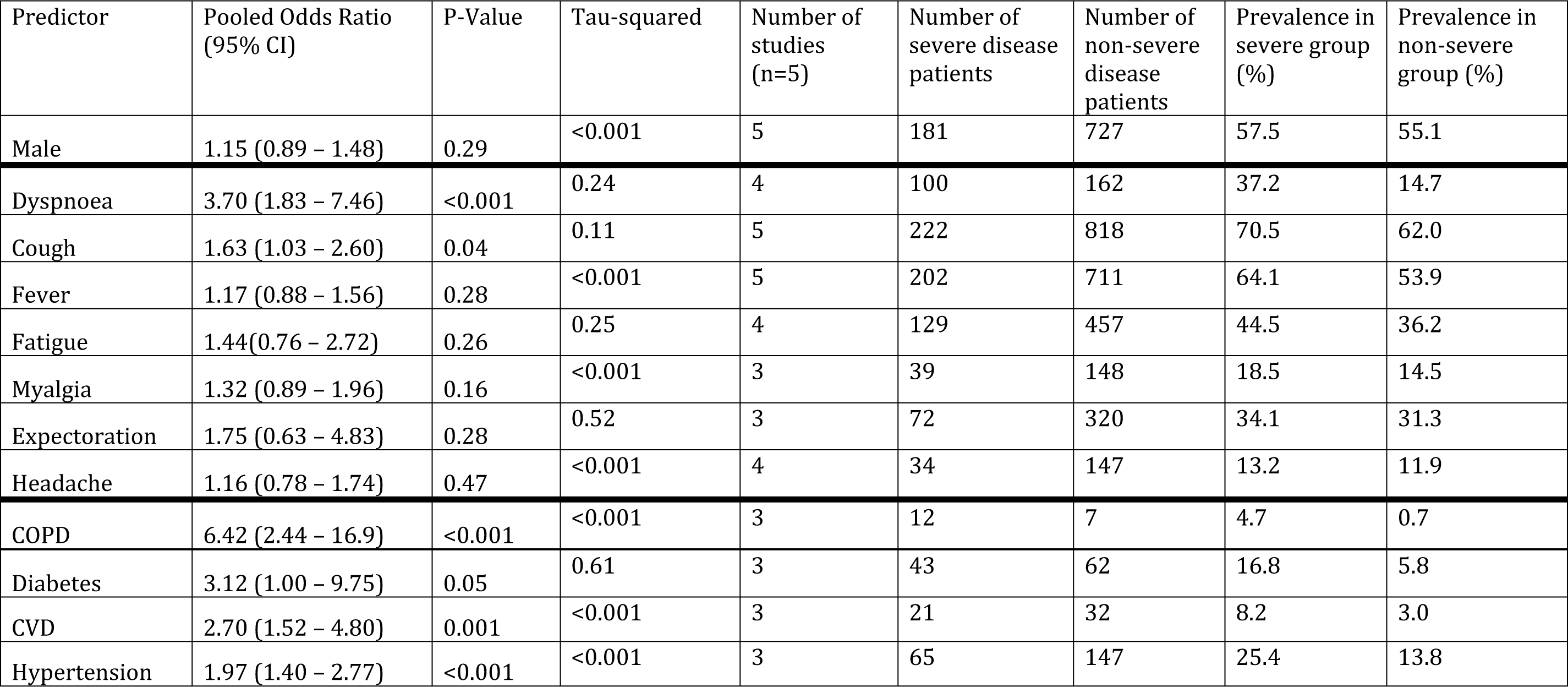
Estimated pOR from meta-analysis for symptoms/comorbidities and severe disease

| Predictor | Pooled Odds Ratio (95% CI) | P-Value | Tau-squared | Number of studies (n=5) | Number of severe disease patients | Number of non-severe disease patients | Prevalence in severe group (%) | Prevalence in non-severe group (%) |
| --- | --- | --- | --- | --- | --- | --- | --- | --- |
| Male | 1.15 (0.89 – 1.48) | 0.29 | <0.001 | 5 | 181 | 727 | 57.5 | 55.1 |
| Dyspnoea | 3.70 (1.83 – 7.46) | <0.001 | 0.24 | 4 | 100 | 162 | 37.2 | 14.7 |
| Cough | 1.63 (1.03 – 2.60) | 0.04 | 0.11 | 5 | 222 | 818 | 70.5 | 62.0 |
| Fever | 1.17 (0.88 – 1.56) | 0.28 | <0.001 | 5 | 202 | 711 | 64.1 | 53.9 |
| Fatigue | 1.44(0.76 – 2.72) | 0.26 | 0.25 | 4 | 129 | 457 | 44.5 | 36.2 |
| Myalgia | 1.32 (0.89 – 1.96) | 0.16 | <0.001 | 3 | 39 | 148 | 18.5 | 14.5 |
| Expectoration | 1.75 (0.63 – 4.83) | 0.28 | 0.52 | 3 | 72 | 320 | 34.1 | 31.3 |
| Headache | 1.16 (0.78 – 1.74) | 0.47 | <0.001 | 4 | 34 | 147 | 13.2 | 11.9 |
| COPD | 6.42 (2.44 – 16.9) | <0.001 | <0.001 | 3 | 12 | 7 | 4.7 | 0.7 |
| Diabetes | 3.12 (1.00 – 9.75) | 0.05 | 0.61 | 3 | 43 | 62 | 16.8 | 5.8 |
| CVD | 2.70 (1.52 – 4.80) | 0.001 | <0.001 | 3 | 21 | 32 | 8.2 | 3.0 |
| Hypertension | 1.97 (1.40 – 2.77) | <0.001 | <0.001 | 3 | 65 | 147 | 25.4 | 13.8 |

**Table 4.**
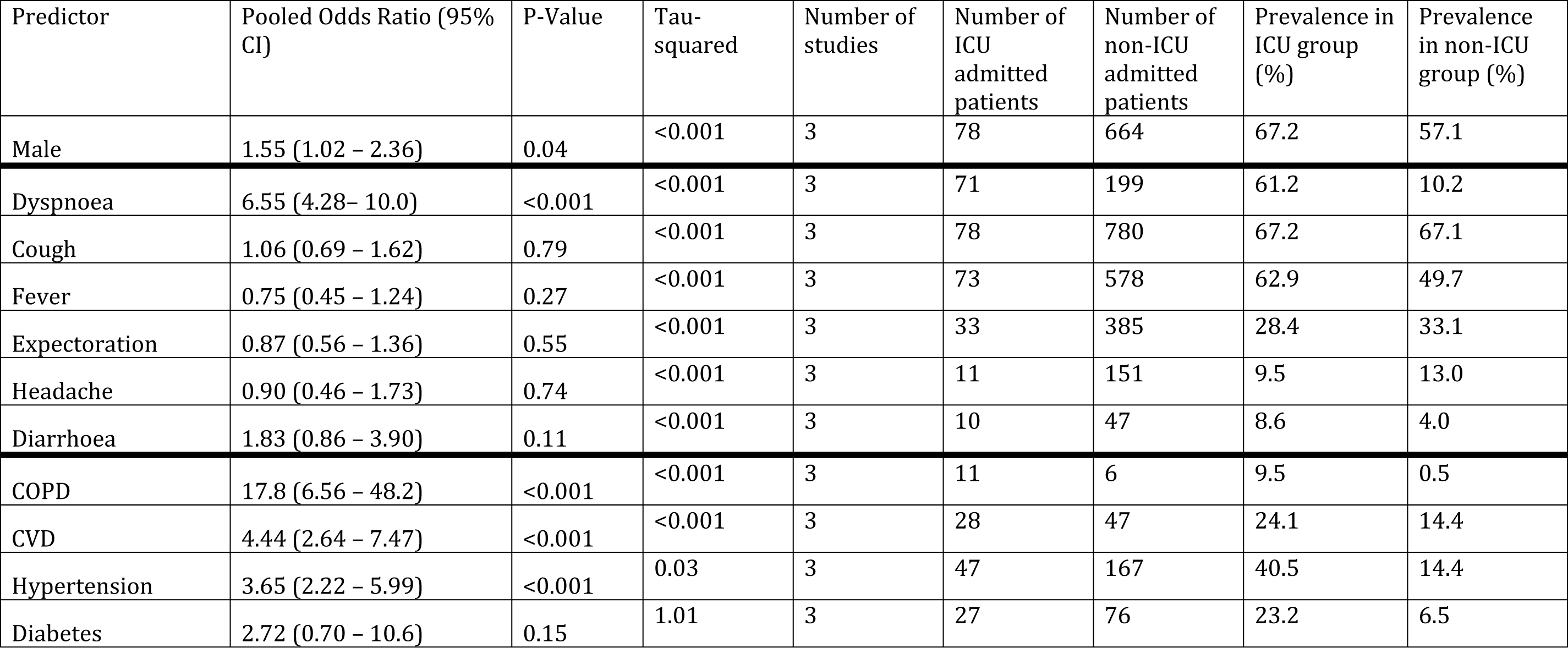
Estimated pOR from meta-analysis for symptoms/comorbidities and ICU admission

Although no more likely to be in the severe group, men were 1.55 times more likely than women to be admitted to ICU (95% CI 1.02 – 2.36). Dyspnoea was the only symptom significantly associated with both severe disease (pOR 3.70, 95% CI 1.83 – 7.46) and ICU admission (pOR 6.55, 95% CI 4.28– 10.0), being more strongly associated with the latter. Cough was associated with severe disease (pOR 1.63, 95% CI 1.03 – 2.60), but not ICU admission. The remaining symptoms analysed were not associated with either outcome. Chronic obstructive pulmonary disease (COPD), cardiovascular disease (CVD), and hypertension were the comorbidities significantly predictive for both severe disease and ICU admission. The pORs for severe disease were as follows: COPD (6.42, 95% CI 2.44-16.9), CVD (2.70, 95% CI 1.52 – 4.80) and hypertension (1.97, 95% CI 1.40 – 2.77). COPD, CVD and hypertension were more strongly associated with ICU admission, compared with severe disease, with pORs of 17.8 (95% CI 6.56 – 48.2), 4.44 (95% CI 2.64 – 7.47), and 3.65 (95% CI 2.22 – 5.99), respectively. In contrast, diabetes was not significantly associated with ICU admission, although the Tau-squared value here was unusually high implying a high level of heterogeneity between studies in this particular case.

## Discussion

### Key Findings

COVID-19 severity was not consistently defined across included studies. All studies were of adequate quality considering the context, and two were of relatively high quality. The ICU group were older (62.4 years) compared to the non-ICU group (46 years), with a significantly higher proportion of males (67.2% vs. 57.1%, p=0.04).

The most prevalent symptoms in the severe disease group were cough (70.5%), fever (64.1%) and fatigue (44.5%), and in the ICU admission group were cough (67.2%), fever (62.9%) and dyspnoea (61.2%). The most prevalent conditions in the severe group were hypertension (25.4%) and diabetes (16.8%), and in the ICU group were hypertension (40.5%) and CVD (24.1%). Males were 1.55 times more likely (95% CI 1.02 - 2.36) to have an ICU admission. Dyspnoea was the only symptom significantly associated with disease severity and ICU admission, alongside various comorbidities (COPD, CVD and hypertension). All of these factors were more strongly associated with ICU admission than disease severity. Patients with dyspnoea were 6.6 times more likely to have an ICU admission (95% CI 4.28– 10.0) compared to those without dyspnoea. Although COPD was relatively uncommon, even in ICU patients, it was by far the most strongly predictive comorbidity for ICU admission (pOR 17.8, 95%CI 6.56 – 48.2). Those with CVD and hypertension were 4.4 (95% CI 2.64 – 7.47) and 3.7 (95% CI 2.22 – 5.99) times more likely to have an ICU admission respectively, compared to patients without the comorbidity.

### Building on existing knowledge

Consistent with Sun et al’s 2020 meta-analysis of symptoms in 50466 COVID-19 patients (5) and the WHO-China joint mission on COVID-19 (18), cough and fever were the most common symptoms found in our analysis. The prevalence of dyspnoea was not investigated in Sun’s meta-analysis, but we found it to be relatively low, particularly in non-severe and non-ICU groups. The prevalence of dyspnoea in the ICU group, however, was 67.2%, compared with 10.2% in the non-ICU group. Whilst dyspnoea is not a particularly common symptom in COVID-19 patients, its significant association with both severe disease and ICU admission may help discriminate between severe and non-severe COVID-19 cases, when present.

The findings reported here are in keeping with current knowledge that the elderly and those with comorbidities are more susceptible to severe infection. (7) According to the Report of the WHO-China Joint Mission on COVID-19 (18), individuals at highest risk for severe disease and death include people with underlying conditions such as hypertension, diabetes, cardiovascular disease, chronic respiratory disease and cancer. We demonstrate that various factors are more strongly associated with ICU admission (representing the very severe end of the disease severity spectrum) compared with less severe disease. This emphasises the extent of the risk COVID-19 poses for patients with comorbidities. The China Centre for Disease Control and Prevention (CDC) found differing case fatality rates (CFRs) in their cohort of 72,314 COVID-19 cases for those with: cardiovascular disease (10.5%), diabetes (7.3%), chronic respiratory disease (6.3%) and hypertension (6.0%)(19). Unlike the China CDC study (19) that presented case fatality rates for different groups, our findings compare those with particular comorbidities to those without, allowing us to estimate the effect of a particular comorbidity on COVID-19 severity. Although we did not investigate death (and included COPD rather than chronic respiratory disease), our analysis similarly suggests that comorbidities are not uniform in terms of the risk of severe COVID-19 disease. Despite being uncommon in our study population, COPD was by far the strongest risk factor for COVID-19 severity, followed by CVD and hypertension.

### Limitations

The foremost limitation of this study was an inability to carry out a multivariable analysis to account for the presence of several symptoms, comorbidities and potential confounders. Although this outbreak has seen the evolution of linked data and large datasets (19) which would be suitable for multivariable analysis, these currently lack the quality of published data: there are large amounts of missing data, a narrow range of collected variables, and uncertainty about data collection methods and consistency. Our univariable analysis is therefore valuable in evaluating specific individual symptoms and comorbidities predictive for COVID-19 severity using high-quality evidence in the form of peer-reviewed studies.

Secondly, the studies included here were all from China, so the generalisability of findings to other countries and populations is not clear. The Chinese may differ to other populations in terms of their health-seeking behaviour, symptom reporting, prevalence of different comorbidities, as well as their access to high quality health services. Nonetheless, given the current dearth of contextually specific evidence available, our findings will help to inform future research and actions in other countries as the outbreak develops.

Finally, it was not possible to account for the timing of presentation in the statistical analysis. If a patient presented after many days of being symptomatic, this may have affected disease severity, compared with an earlier clinical presentation. However, this limitation does not apply to comorbidities, and Table 2 shows information from individual studies on median duration of symptoms before admission, which appears similar between severe (or ICU) and non-severe (or non-ICU) cases. It is therefore unlikely that this will have biased the overall results.

### Implications for clinical practice/public health

By identifying the symptoms and comorbidities predictive for more severe disease, clinicians can better stratify the risk of individual patients, as early as their initial contact with health services. This can lead to practical changes in management, which can improve allocative efficiency as well as clinical outcomes, through the consideration of more intensive environments of care (e.g. high dependency unit), earlier on, for patients at highest risk of severe infection. These can also be formalised within risk stratification tools to aid clinical decision-making, such as the CURB65 tool for community-acquired pneumonia (20). As the number of hospitalised COVID-19 cases continues to increase, hospitals will increasingly need to ration limited resources and improve clinical pathways to effectively prioritise patients with greatest clinical need. This is important, as COVID-19 is already placing increased pressure on ICUs, and anticipation of future demand, based on local population characteristics, may enable more timely planning and resource mobilisation (21). Identifying those at the highest risk will also facilitate better-informed discussions between clinicians, patients and patients’ families about the anticipated clinical trajectory, allowing more accurate and timely advance care discussions to occur.

Identifying those at high-risk will aid the public health response in controlling the spread of disease. Given the ubiquity of comorbidities in the elderly population, and their increased susceptibility to severe COVID-19 infection (18), knowledge on the differing prevalence and risk of various conditions may help to focus and tailor public health efforts such as the screening of asymptomatic individuals, risk communication, contact tracing, self-isolation and social distancing. For instance, for COPD, which is less common in the general population and very strongly associated with ICU admission, a more targeted and intensive health protection strategy may be warranted, compared to other conditions (such as hypertension) that are more difficult to target due to their higher prevalence in the general population.

Furthermore, if it is found that severity of illness is related to infectivity, as is the case in the closely related SARS-CoV, then identifying patients who may develop severe illness can help guide precautions to prevent the spread of SARS-CoV2. These include infection control decisions regarding the limited availability of isolation rooms and personal protective equipment (PPE), particularly in more resource-constrained settings. This will be of particular importance as the outbreak develops, if the prevalence of hospitalised COVID-19 patients increases.

### Implications for future research

Measurement tools used in studies should aim to objectively measure symptoms where possible, and details on how and when symptoms were ascertained should be made clear in all studies. There was also heterogeneity in the range of symptoms and comorbidities recorded by different studies, and some studies grouped various symptoms (8-10), limiting the utility of such data.

Studies investigating COVID-19 severity should avoid using subjective or self-reported criteria (such as dyspnoea) in the definition of severity, as in one study included here (13). To ensure consistency between studies and comparability, the WHO-China Joint Mission on COVID-19 definition of severe and critical cases should be used. Furthermore, the distinction between cases that are severe and critical should be clearly made in studies, to enable more accurate risk stratification of the most unwell patients.

For future research on predictors of severity, research should aim to include greater detail on specific conditions, including how well controlled chronic conditions were before and during admission. If the severity of COVID-19 varies according to the severity of underlying comorbidities, there may be a case for optimising routine treatment for healthy, uninfected individuals, as a potential public health action to mitigate risk. Multivariable analysis to identify which groups of symptoms or comorbidities are most associated with severe or critical disease will also be valuable.

### Conclusions

The existing literature on COVID-19 fails to elucidate the specific symptoms and comorbidities most predictive for severe COVID-19 cases. Our analysis finds that dyspnoea is the only symptom strongly predictive for both severe disease and ICU admission, and could be a useful symptom to help guide clinical management decisions early in the course of illness. The association between comorbidities and severe disease is not homogenous. Whilst COPD, cardiovascular disease, and hypertension were all associated with severity, COPD was the most strongly predictive. When looking at ICU-admitted patients, who represent the more severe end of the spectrum of clinical severity, the difference in effect sizes for COPD and the other included comorbidities was large, suggesting COPD patients are particularly vulnerable to critically severe disease. As the outbreak develops, future research must aim to substantiate these findings by investigating factors related to disease severity. This will aid clinical assessment, risk stratification, and resource allocation, and allow public health interventions to be targeted at the most vulnerable.

## Data Availability

All data are fully available without restriction

